# Immunohistochemical expression of TFF1 is a new prognostic marker in retinoblastoma

**DOI:** 10.1101/2023.07.24.23293064

**Authors:** Rosario Aschero, Daiana Ganiewich, Gabriela Lamas, Camilo A Restrepo-Perdomo, Daniela Ottaviani, Santiago Zugbi, Sandra Camarero, Ezequiel Néspoli, Maria Cuadrado Vilanova, Sara Perez-Jaume, Guillem Pascual-Pasto, Claudia Sampor, Nathalia Grigorovski, Beatriz Salas, Mariona Suñol, Angel M. Carcaboso, Jaume Mora, María T G de Dávila, François Doz, François Radvanyi, David H Abramson, Andrea S Llera, Paula S Schaiquevich, Fabiana Lubieniecki, Guillermo L Chantada

**Affiliations:** Pathology Service, Hospital de Pediatría JP Garrahan, Buenos Aires 1245, Argentina; National Scientific and Technical Research Council, CONICET, Buenos Aires 1425, Argentina; SJD Pediatric Cancer Center Barcelona, Hospital Sant Joan de Deu, Barcelona, 08950, Spain; Institut de Recerca Sant Joan de Deu, Barcelona, 08950, Spain; Instituto de Investigaciones en Medicina Traslacional - Universidad Austral, Buenos Aires, Argentina; Pathology Service, Hospital Sant Joan de Deu, Barcelona, 08950, Spain; SIREDO Center, Institut Curie and University Paris Cité, Paris, France; Unidad de tratamientos innovadores, Hospital de Pediatría JP Garrahan, Buenos Aires 1245, Argentina; Hematology-Oncology Service, Hospital de Pediatría JP Garrahan, Buenos Aires 1245, Argentina; Department of Pediatric Oncology, Clinical Division, National Institute of Cancer, Rio de Janeiro, Brazil; Hospital del Niño Manuel A. Villarroel, Cochabamba 2500, Bolivia; Ophthalmic Oncology Service, Memorial Sloan-Kettering Cancer Center, NY 10065, US; Laboratory of Molecular and Cellular Therapy, Instituto Leloir-Instituto de Investigaciones Bioquímicas de Buenos Aires (IIBBA), Buenos Aires 1405, Argentina; Hematology Oncology Service, Hospital Pereyra Rossell, Montevideo, Uruguay

**Keywords:** High-risk retinoblastoma, Prognostic marker, TFF1 expression, Histological subtypes

## Abstract

**Introduction:** The risk of relapse in retinoblastoma is currently determined by the presence of high-risk histopathologic factors in the enucleated eye. However, the probability of developing metastatic disease is heterogeneous among these patients. Evaluating a biological marker to identify high-risk patients could be useful in clinical setting. This study aims to evaluate whether the expression of TFF1, a surrogate for subtype 2 retinoblastoma, is a prognostic marker for relapse and death.

**Methods:** This multicenter cohort study included 273 patients, 48 of whom had extraocular disease. Immunohistochemical staining were performed for CRX, ARR3, TFF1 and Ki67. Tumors were classified as histological subtype 1 (HS1) if they had low or no expression of TFF1 (quick score (QS) ≤ 50) and as histological subtype 2 (HS2) if they expressed TFF1 diffusely (QS > 50). We studied the association between HS classification and outcome.

**Results:** Of 273 patients, 35.9% were classified as HS1, 59.3% as HS2 and 4.8% were not evaluable. In multivariate analysis, patients with HS2 tumors had a higher probability of relapse and death than those with HS1 (*P* < 0.0001 and *P* = 0.00020, respectively). We identified a higher-risk subgroup among HS2 tumors, presenting non-mutually exclusive expression of ARR3 and TFF1 and had an increased risk of relapse and death compared to tumors that displayed mutually exclusive expression (*P* = 0.012 and *P* = 0.027, respectively).

**Conclusions:** Expression of TFF1, especially when it is not-mutually exclusive with ARR3, is an independent prognostic marker of poor outcome in retinoblastoma.

## INTRODUCTION

Retinoblastoma is the most common pediatric ocular cancer and one of the index tumors proposed by the World Health Organization for global action^1^. Though highly curable in high income countries, worldwide about 50% of affected children die from metastases^2^.

Many patients need enucleation of the affected eye for treatment and the most important high-risk histopathologic factors (HRPF) for extraocular relapse are invasion of choroid, optic nerve and sclera^3^. Tumors without these features are very unlikely to develop extraocular disease, and no further treatment after enucleation is recommended^4^. Although HRPF correlate with a greater risk of metastases, most children whose tumors display these features will never develop metastatic relapse^5,6^. There is no clear consensus about who should receive adjuvant therapy to reduce the risk of metastatic relapse since this risk is variable among the different groups^6-8^. Furthermore, in addition to the presence of HRPF, there may be undefined biological features that determine the risk of relapse.

Recently, two retinoblastoma biological subtypes have been described by multi-omic analysis. Subtype 1 tumors express genes associated with late cone differentiation, have few genetic alterations other than *RB1* inactivation. In contrast, subtype 2 tumors exhibit stemness features, intratumoral heterogeneity expressing less differentiated cone and neuronal/ganglion cell markers and nearly all harbor other genetic alterations^9^. Since metastatic dissemination was reported in subtype 2 but not in subtype 1 tumors, it would be critical to identify subtype 2 tumors in clinical settings mainly where multi-omic analysis are not widely available, like in countries with limited resources where HRPF are more prevalent.

In subtype 2 tumors, gene expression analysis showed that the *Trefoil factor 1* (*TFF1*), is the top up-regulated gene^9^. TFF1 is not expressed in the normal retina^10^ and has little or no expression in subtype 1 tumors^9^. TFF1 antibodies are commercially available for immunohistochemistry, that might be useful for identifying subtype 2 tumors in the clinical setting.

In the present study, we aimed to evaluate the expression of TFF1 by immunohistochemistry, as histological surrogate for subtype 2 retinoblastoma and assess its potential role as a predictive risk factor for metastatic relapse and survival in a large series of patients.

## METHODS

### Patients

We included an initial cohort of 252 consecutive patients with retinoblastoma stage 0-I with HRPF and II-IV according to the International retinoblastoma staging system (IRSS)^11^ from Hospital JP Garrahan (HPG, Argentina) treated between 1988 and 2020 and all enucleated eyes regardless of HRPF from Hospital Sant Joan de Déu (SJD, Spain) between 1983 and 2018.

The inclusion criteria were the availability of biological samples that were clinically annotated. Exclusion criteria was the diagnosis of trilateral disease because the outcome is not dependent on HRPF.

To increase the number of cases with extraocular disease, we included an expanded cohort of patients with extraocular disease from other collaborating institutions. However, since these patients were not consecutive, they were included only in immunohistochemistry descriptive analysis and were excluded of all statistical analyses. Therefore, a total of 273 patients were included in the study.

### Patient classification

Patients were classified according to the IRSS (**Figure S1**). Patients from the initial cohort with intraocular disease at diagnosis (*N* = 242) were either secondary enucleated after eye-conservative treatment (*N* = 28), enucleated as first-line treatment (*N* = 206), or treated with neoadjuvant therapy because of buphthalmos (*N* = 5)^12^. In three cases, parents declined enucleation when it was indicated, and these patients were later enucleated. Ten children presented with extraocular disease at diagnosis, of whom nine received neoadjuvant therapy prior to enucleation.

The expanded cohort (*N* = 21) included additional patients with disseminated retinoblastoma. Fourteen patients had intraocular disease at diagnosis, five had extraocular retinoblastoma, and two children developed extraocular disease due to enucleation refusal at diagnosis.

### Clinical and histopathological studies

For each patient we retrieved clinical, treatment and survival data from each institutional database. For the histological study, pathology slides were blindly reviewed by two independent researchers and tumors were retrospectively assigned a pTNM stage^13^ when not recorded prospectively.

Immunohistochemical staining was performed on 3 µm thick sections of paraffin-embedded tissue, selecting, when available, the block with the central pupil-optic nerve section, ideally containing the optic nerve, tumor and anterior chamber structures^14^. To characterize the expression of photoreceptor-associated markers, we evaluated the cone-rod homeobox (CRX) (ab140603; Abcam, Cambridge, UK), an early photoreceptor marker and arrestin-C (ARR3) (11100-2-AP; Proteintech Group, Manchester, UK) as a marker of late cone differentiation. Expression of TFF1 (HPA003425; Sigma-Aldrich Sigma, St. Louis, USA) was selected as a surrogate marker for subtype 2 tumors. We also evaluated Ki67 expression in all cases to assess the cell proliferation index (M7240, Agilent, Santa Clara, USA). We revealed the slides using Novolink Polymer detection System (Leica Biosystems, Wetzlar, Germany).

Additional information about the conditions and positive controls used are described in **Table S1**. To quantify the immunostaining results, the quick score (QS) was performed as reported previously^9,15^. Briefly, the intensity was classified as negative (0), mild (1), moderate (2) or strong (3) and the percentage of positive cells at any intensity level was calculated. The QS was calculated as (Intensity^highest^)*(Percentage^total^), and the result score from 0 to 300. QS value reported is the mean of QS resulting from the assessment of two specialist. In case of discrepancy, a third observer evaluated the slides and the case was classified by consensus. Using previously reported criteria, we defined histological subtype 1 (HS1) tumors as those with a TFF1 QS value ≤ 50 and histological subtype 2 (HS2) tumors as those with TFF1 QS value > 50^9^ in enucleated eyes or metastatic sites when no ocular tissue was available. Patients with bilateral disease enucleated from both eyes who presented a different subtype in each eye were classified as HS2 (**Table S2**).

### Statistical analyses

Statistical analyses were performed in R/RStudio v4.1.0. Comparisons of clinical and histological features were performed using the Wilcoxon test for continuous variables and the Fisheŕs exact test for categorical variables. A *P* value cut-off of 0.05 was used to assess statistical significance. Event-free survival (EFS) and overall survival (OS) were defined as the time from diagnosis to first event (relapse or death, respectively) or last follow-up. Kaplan-Meier^16^ plots and log-rank test^17^ were used to compare the survival distributions (GraphPad Prism 8 Software, La Jolla, CA, USA). Cox proportional hazards regression models^18^ with Firth correction^19^ were used to test associated with outcome. Only variables statistically significant in the univariate analysis were included in the multivariate analysis.

For survival analysis, patients who refused enucleation were censored and those with non-evaluable HRPF or HS and one who developed a second tumor were excluded from these analyses.

### Ethics statements

The HPG Institutional Review Board approved this study (Protocol #979). The study was conducted under the tenets of the Declaration of Helsinki.

## RESULTS

### Patients characteristics

Clinical and histopathological characteristics are listed in **Table S3** and **Table S4** respectively. Of the 271 enucleated eyes, 181 (66.3%) presented HRPF. All patients who refused eye enucleation at diagnosis and were later enucleated (5/5) had HRPF. In six tumors (2.2%) from patients receiving neoadjuvant therapy, the presence of HPRF was not evaluable due to necrosis greater than 90%.

### Immunohistochemistry

All tumors included in the study were positive for CRX nuclear staining consistent with the cone origin of human retinoblastoma. ARR3 expression was positive in all eyes in most tumor cells although at different intensities. The strongest expression was found in the most differentiated tumor cells, consistent with ARR3 expression at later stages of the cone cell differentiation process. Expression of TFF1 was cytoplasmic, of heterogeneous intensity and displayed high inter- and intra-tumoral variability.

Based on the TFF1 expression, of 273 patients evaluated, 98 (35.9%) were classified as HS1 and 162 (59.3%) as HS2. In twenty-two of 273 (8.1%) patients it was not possible to assign a HS by examination of the enucleated eye because of necrosis, but in nine of them it could be assigned by evaluating a biopsy of a metastatic site (**Table S2**). Therefore, 13 (4.8%) patients remain as not evaluable. Examples of each marker immunohistochemistry are presented in **Figure 1A**. Overall, 24 specimens of metastatic sites were available for histological evaluation, 22 (91.7%) were assigned to HS2 and two (8.3%) to HS1 corresponding to one patient whose family refused enucleation and one child with orbital extension at diagnosis who achieved long-term survival (**Figure 1B**).

**Figure 1.**
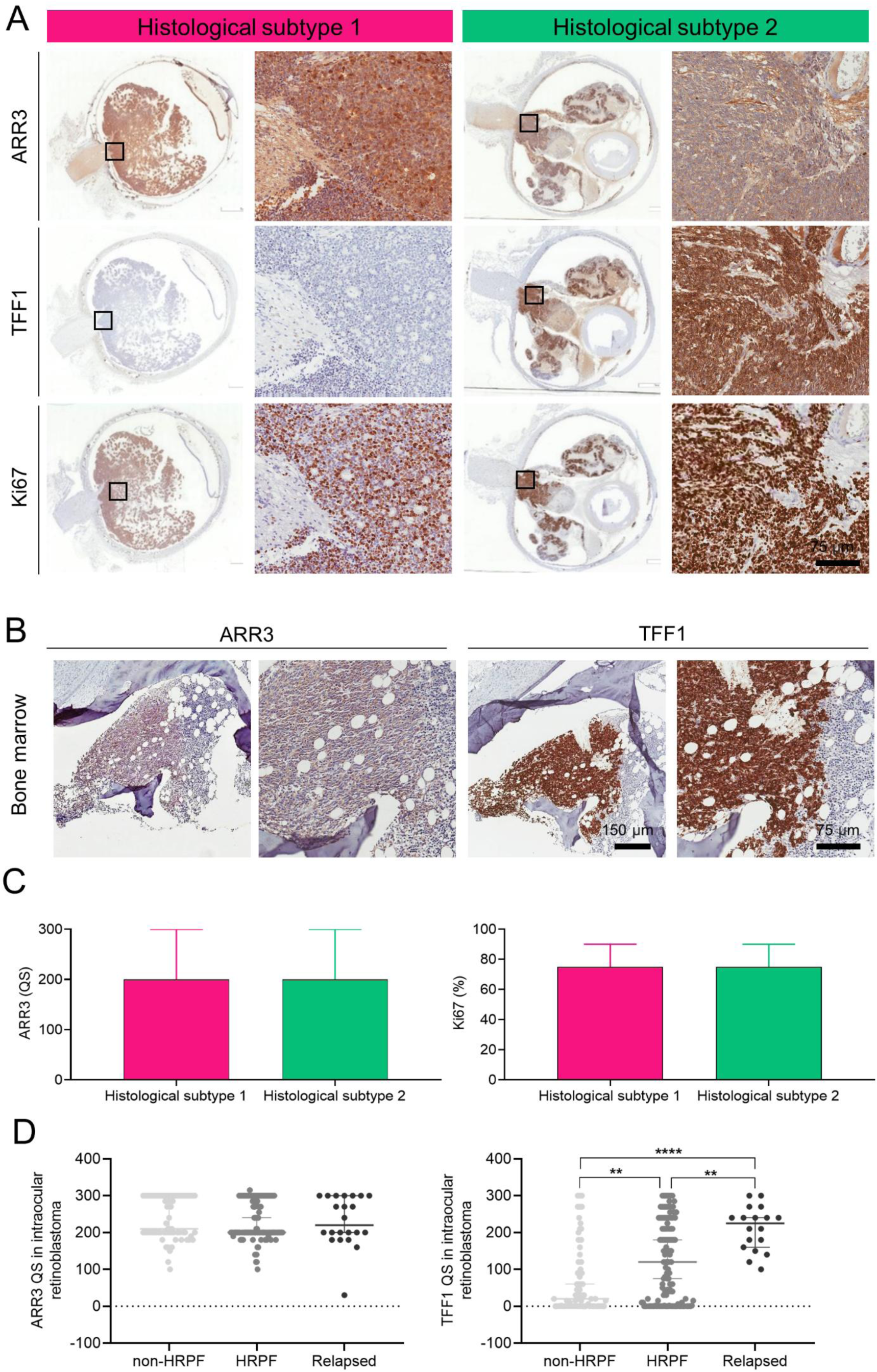
Immunohistochemical panel expression. Expression of antibodies included in the immunohistochemical panel in HS1 and HS2 tumors. (A) HS1 tumors were negative or focally positive (QS ≤ 50) for TFF1 expression, while tumors classified as HS2 were positive (QS > 50) (Patients RB218 and RB152, respectively). Representative images at panoramic and 4x magnification. (B) Immunostaining of ARR3 and TFF1 in bone marrow metastasis (patient RB040) at 4x and 10x magnification. Retinoblastoma cells were positive for both markers. (C) Median values with interquartile range of QS and percentage of positive cells, obtained for the differentiation marker ARR3 and Ki67, respectively. Mann-Whitney test was used. (D) TFF1 and ARR3 QS of patients with intraocular disease at diagnosis included in the initial cohort, grouped in patients with or without risk factors (HRPF and non-HRPF, respectively) that did not develop metastasis, and patients that developed metastasis (relapsed). Kruskal-Wallis and Dunńs multiple comparisons tests were used. \*\**P* ≤ 0.01 and \*\*\*\**P* ≤ 0.0001.

The expression of ARR3 was similar in HS1 (median QS = 200, range 100 to 300) to HS2 (median QS = 200, range 30 to 300) (*P* = 0.8953). The percentage of positive cells for Ki67 was similar in HS1 (median = 75%, range 10% to 95%) compared to tumors classified as HS2 (median = 75%, range 40% to 99%) (*P* = 0.3854) (**Figure 1C**).

The TFF1 QS values for tumors that developed metastatic relapse (median = 225, range 100 to 300) were significantly higher compared to those of patients with intraocular tumors that did not develop metastasis, including cases with (median = 120, range 0 to 300) or without HRPF (median = 25, range 0 to 300) (*P* < 0.0001). This positive association was not found for ARR3 (*P* = 0.8757) (**Figure 1D)**.

### Clinico-pathological correlations

Patients from the initial cohort with intraocular retinoblastoma and HS1 tumors were significantly younger at diagnosis compared with HS2 tumors. By contrast, children with intraocular disease at diagnosis and HS2 tumors presented more frequently HRPF in the enucleated eyes, had significantly higher risk of extraocular relapse and death due to progression of disease compared to HS1 tumors (**Table 1**).

**Table 1.**
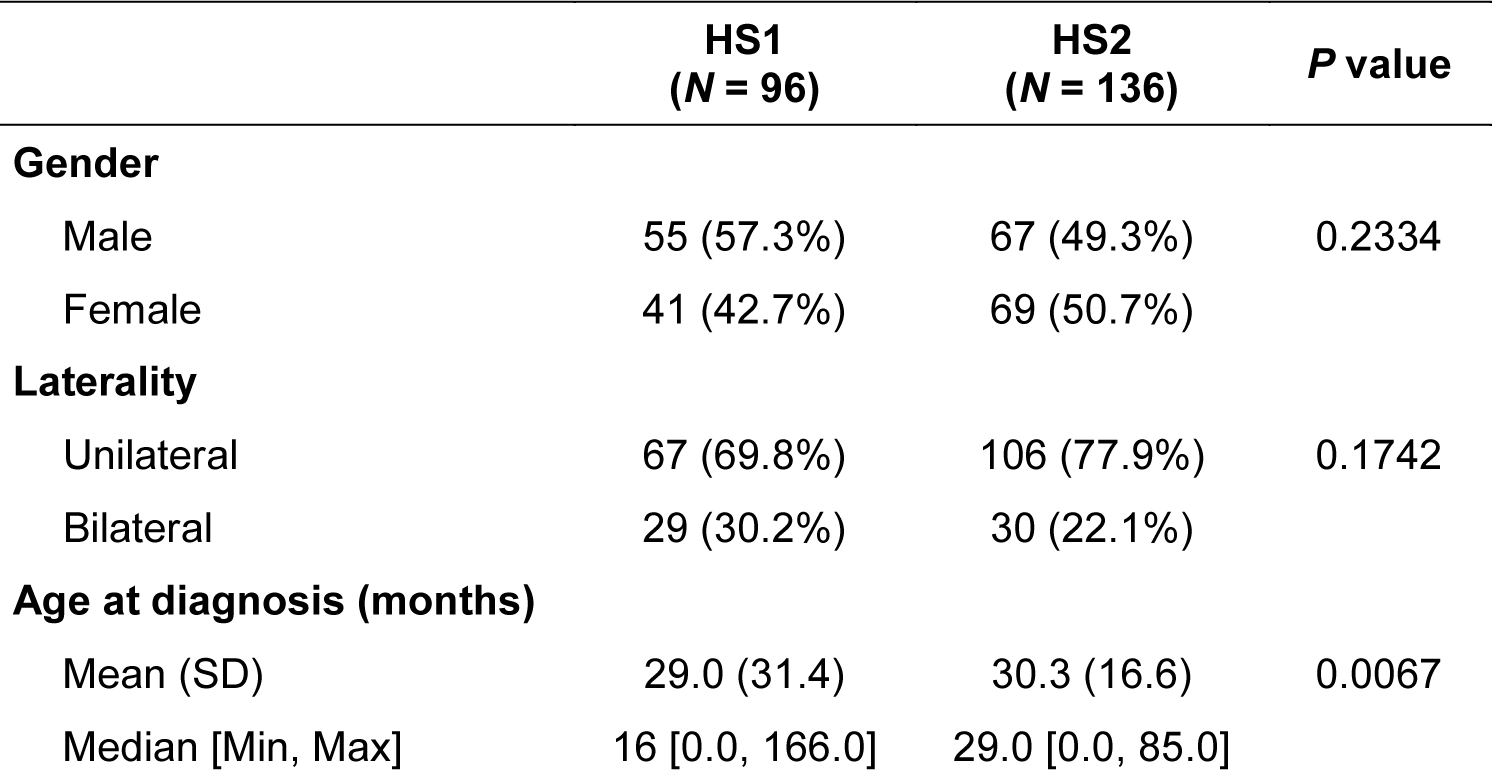
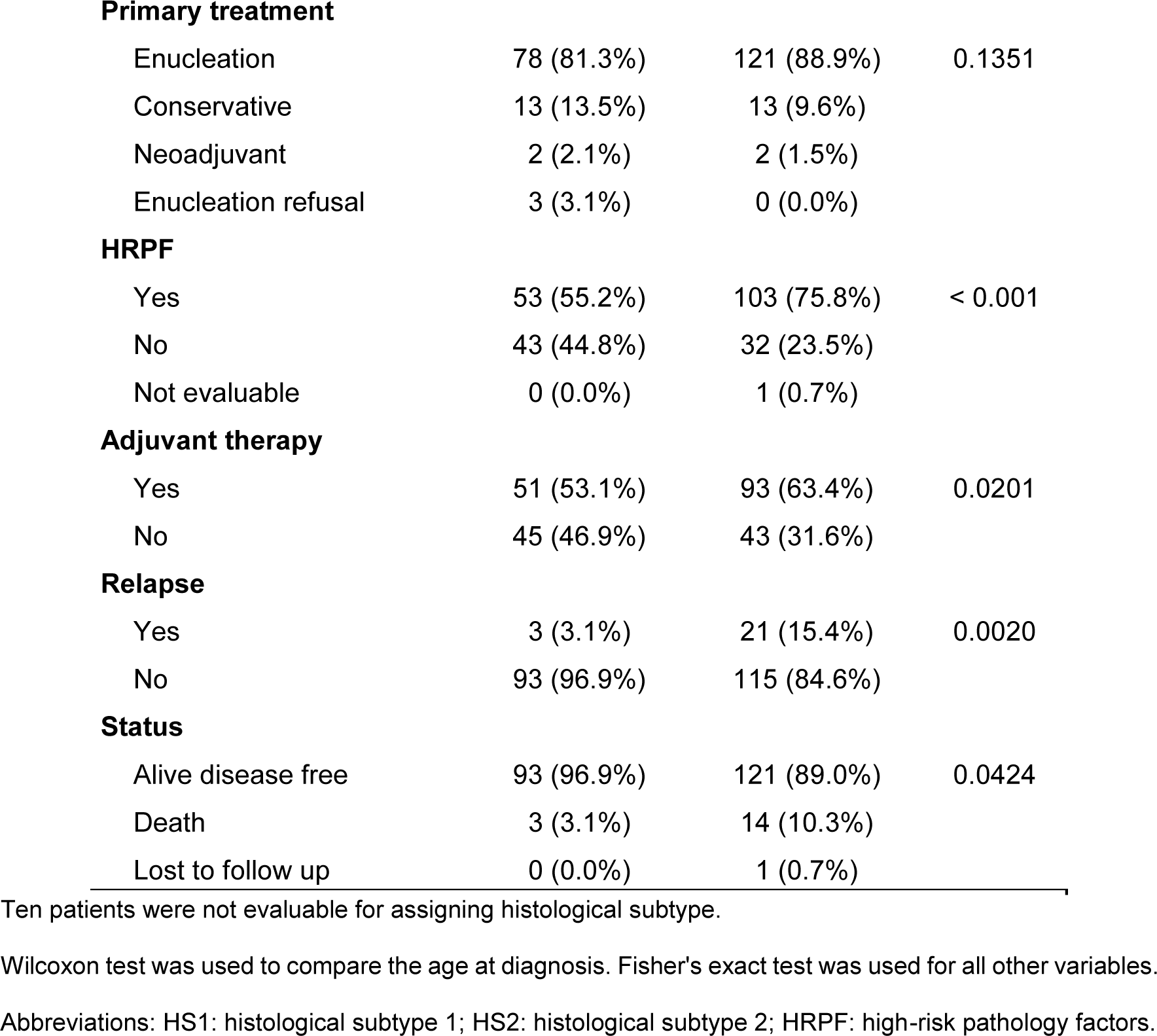
Clinical characteristics of both histological subtype tumors in patients with intraocular disease included in the initial cohort (*N* = 242).

### Extraocular relapse according to histological subtype

Twenty-four patients (24/242, 9.9%) with intraocular disease of the initial cohort had an extraocular relapse after enucleation. Clinical characteristics of this subgroup are detailed in **Figure S2**.

Overall, in the initial cohort, 228 (90.5%) patients are alive and disease free with a median follow-up of 62 months (range 2 to 370) and 23 (9.1%) died of disease with a median survival of 15 months (range 1 to 82).

Five-year event free survival and OS for patients with HS1 tumors was significantly higher than compared to patients with HS2 tumors (*P* = 0.0003 and *P* = 0.0019, respectively) (**Figure 2A**). The 5-year EFS using HS classification were 100.0% (95% CI 100.0% to 100.0%) for HS1 and 85.3% (95%CI 79.2% to 91.9%) for HS2, whilst the 5-year OS were 100.0% (95% CI 100.0% to 100.0%) and 88.8% (95% CI 83.2 to 94.8%), respectively. Multivariate Cox analysis showed a significantly higher risk of extraocular relapse and death for patients with HS2, and tumors with postlaminar infiltration of the optic nerve. **Table 2** summarizes the statistical significance of the main variables. All factors evaluated in univariate analysis and their results are listed in **Table S5** and **Table S6.**

**Figure 2.**
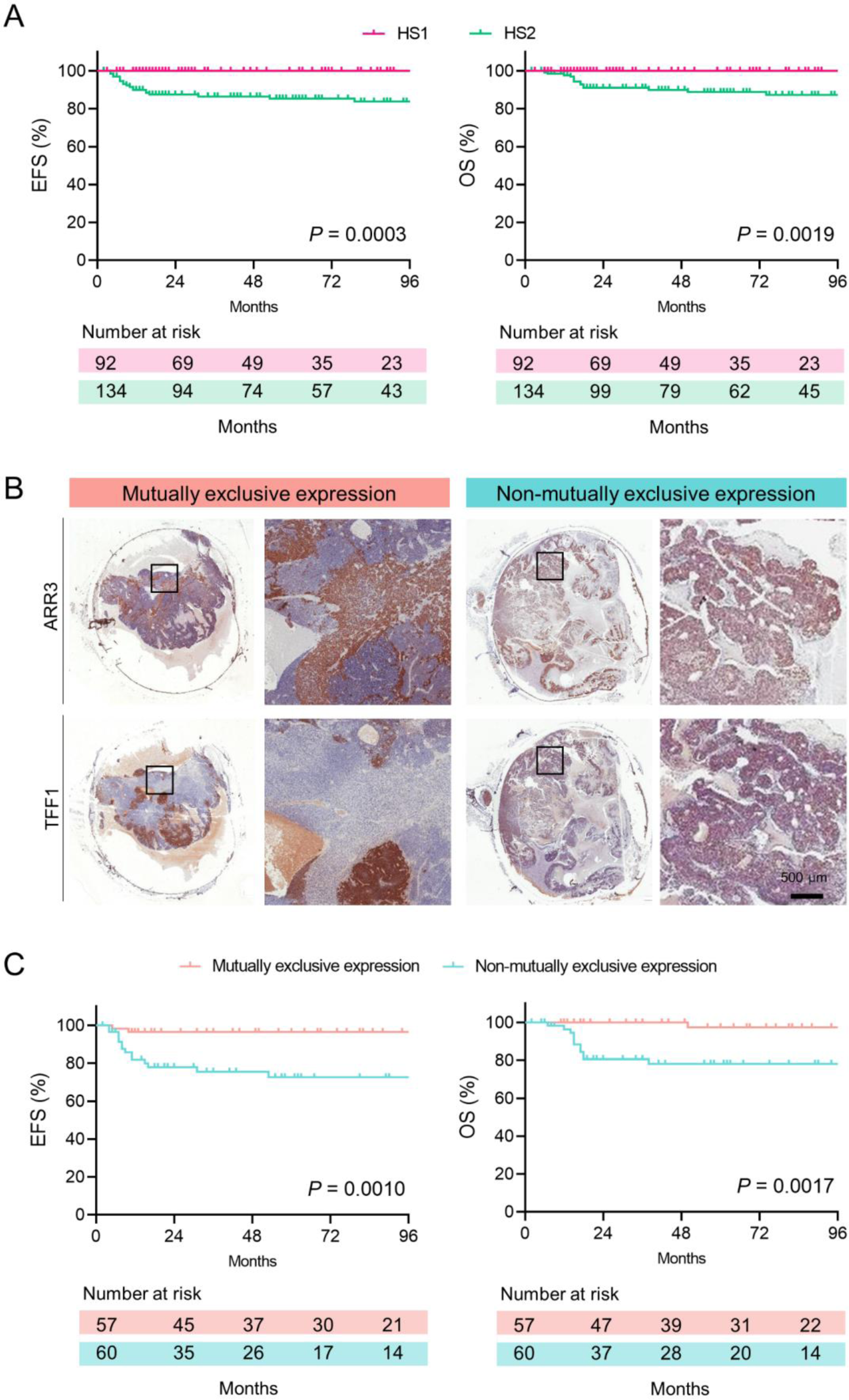
Histological subtype 2 tumors are associated with higher risk of relapse. (A) Event-free survival and OS of patients included in the initial cohort with intraocular disease at diagnosis stratified according to the histological subtype classification. (B) Representative images for expression of ARR3 and TFF1 in mutually and non-mutually exclusive expression patterns (Patient RB098 and RB011 respectively) at panoramic and 2x magnification. Histological subtype 2 tumors display intratumoral heterogeneity, with areas ARR3+/TFF1- intertwined with ARR3-/TFF1+ regions (*N* = 57) or ARR3+/TFF1+ in all tumor cells (*N* = 60). (C) Event-free and overall survival of patients included in the initial cohort with intraocular disease and HS2, classified according to the presence or absence of the dual expression pattern.

**Table 2.**
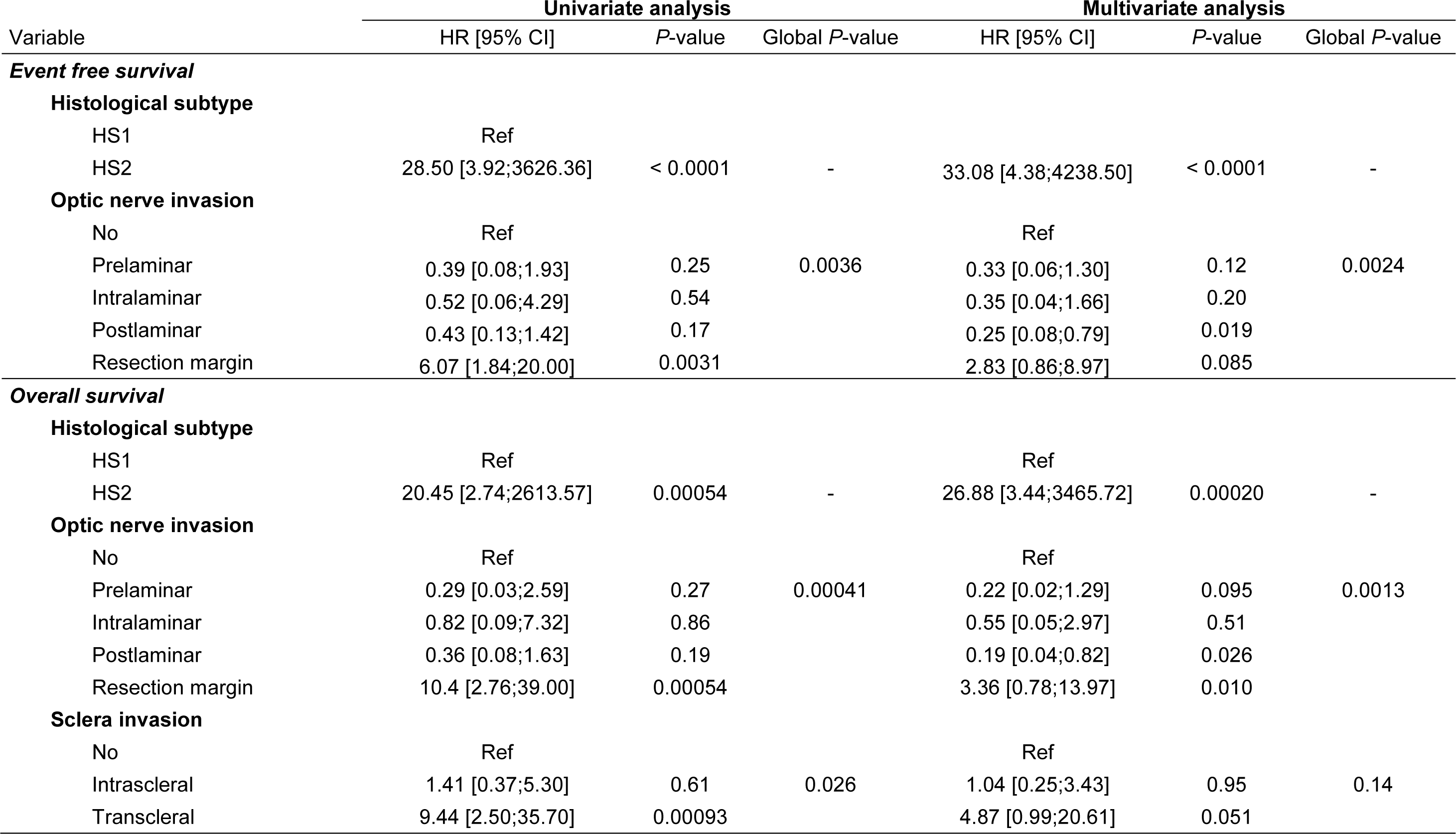
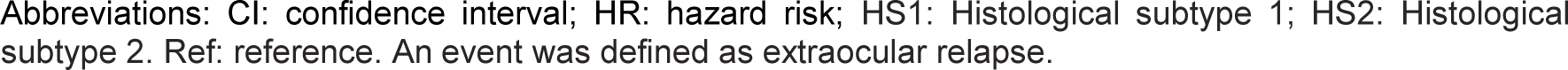
Cox proportional hazards regression models in retinoblastoma patients for association of retinoblastoma features and histological subtypes classification with probability of event-free survival and overall survival.

| Variable | Univariate analysis |  |  | Multivariate analysis |  |  |
| --- | --- | --- | --- | --- | --- | --- |
|  | HR [95% CI] | P-value | Global P-value | HR [95% CI] | P-value | Global P-value |
| **Event free survival** |  |  |  |  |  |  |
| **Histological subtype** |  |  |  |  |  |  |
| HS1 | Ref |  |  |  |  |  |
| HS2 | 28.50 [3.92;3626.36] | < 0.0001 | - | 33.08 [4.38;4238.50] | < 0.0001 | - |
| **Optic nerve invasion** |  |  |  |  |  |  |
| No | Ref |  |  | Ref |  |  |
| Prelaminar | 0.39 [0.08;1.93] | 0.25 | 0.0036 | 0.33 [0.06;1.30] | 0.12 | 0.0024 |
| Intralaminar | 0.52 [0.06;4.29] | 0.54 |  | 0.35 [0.04;1.66] | 0.20 |  |
| Postlaminar | 0.43 [0.13;1.42] | 0.17 |  | 0.25 [0.08;0.79] | 0.019 |  |
| Resection margin | 6.07 [1.84;20.00] | 0.0031 |  | 2.83 [0.86;8.97] | 0.085 |  |
| **Overall survival** |  |  |  |  |  |  |
| **Histological subtype** |  |  |  |  |  |  |
| HS1 | Ref |  |  | Ref |  |  |
| HS2 | 20.45 [2.74;2613.57] | 0.00054 | - | 26.88 [3.44;3465.72] | 0.00020 | - |
| **Optic nerve invasion** |  |  |  |  |  |  |
| No | Ref |  |  | Ref |  |  |
| Prelaminar | 0.29 [0.03;2.59] | 0.27 | 0.00041 | 0.22 [0.02;1.29] | 0.095 | 0.0013 |
| Intralaminar | 0.82 [0.09;7.32] | 0.86 |  | 0.55 [0.05;2.97] | 0.51 |  |
| Postlaminar | 0.36 [0.08;1.63] | 0.19 |  | 0.19 [0.04;0.82] | 0.026 |  |
| Resection margin | 10.4 [2.76;39.00] | 0.00054 |  | 3.36 [0.78;13.97] | 0.010 |  |
| **Sclera invasion** |  |  |  |  |  |  |
| No | Ref |  |  | Ref |  |  |
| Intrascleral | 1.41 [0.37;5.30] | 0.61 | 0.026 | 1.04 [0.25;3.43] | 0.95 | 0.14 |
| Transcleral | 9.44 [2.50;35.70] | 0.00093 |  | 4.87 [0.99;20.61] | 0.051 |  |

Abbreviations: CI: confidence interval; HR: hazard risk; HS1: Histological subtype 1; HS2: Histological subtype 2. Ref: reference. An event was defined as extraocular relapse.

### Risk factors for extraocular relapse in HS2

Since all cases with extraocular relapse (except those that occurred in patients whose families declined enucleation) occurred in HS2, we analyzed this cohort separately to further screen for those with the highest risk of relapse. As previously reported for EBF3 (another marker for subtype 2 tumors)^9^, we found two different patterns of expression of TFF1 and ARR3: (i) a non-mutually exclusive expression characterized by the expression of ARR3 and TFF1 (ARR3+/TFF1+) in all tumor cells, shown in 44.1% (60/136) of samples; and (ii) a mutually exclusive expression pattern whereby areas of ARR3+/TFF1- cells were alternated with regions ARR3-/TFF1+ seen in 41.9% (57/136) of cases. In the mutually exclusive pattern, the ARR3+/TFF1- area corresponded to well-differentiated tumor cells with the presence of rosettes in 77.2% (44/57) of cases (**Figure 2B**). In 13.9% (18/136) of samples the dual pattern of expression could not be determined, and these cases were excluded from the analysis.

Patients with HS2 tumors with a mutually exclusive expression were significantly younger at diagnosis compared to patients with the non-mutually exclusive expression pattern (median 24.5 months, range 0 to 85 vs 32.5 months, range 1 to 82 respectively; *P* = 0.0035). The non-mutually exclusive expression pattern was associated with a higher risk of relapse and death of disease compared to tumors with the mutually exclusive expression pattern ARR3+/TFF1- or ARR3-/TFF1+ (*P* = 0.0010 and *P* = 0.0017, respectively) (**Figure 2C**). The 5-year EFS and OS were 96.5% (95% CI 91.8% to 100.0%) and 97.4% (95% CI 92.4% to 100.0%) for HS2 mutually exclusive expression tumors, and 72.6% (CI 95% 61.0% to 86.0%) and 78.1% (95% CI 67.4% to 90.6%) for HS2 non-mutually exclusive expression pattern, respectively.

In the multivariate analysis performed in HS2 tumors from the initial cohort, the non-mutually exclusive expression pattern was the only statistically significant predictor of EFS, whilst together with postlaminar invasion of the optic nerve remained significant in the OS analysis (**Table 3**). All studied factors included in the univariate analysis are shown in **Table S7** and **Table S8**.

**Table 3.**
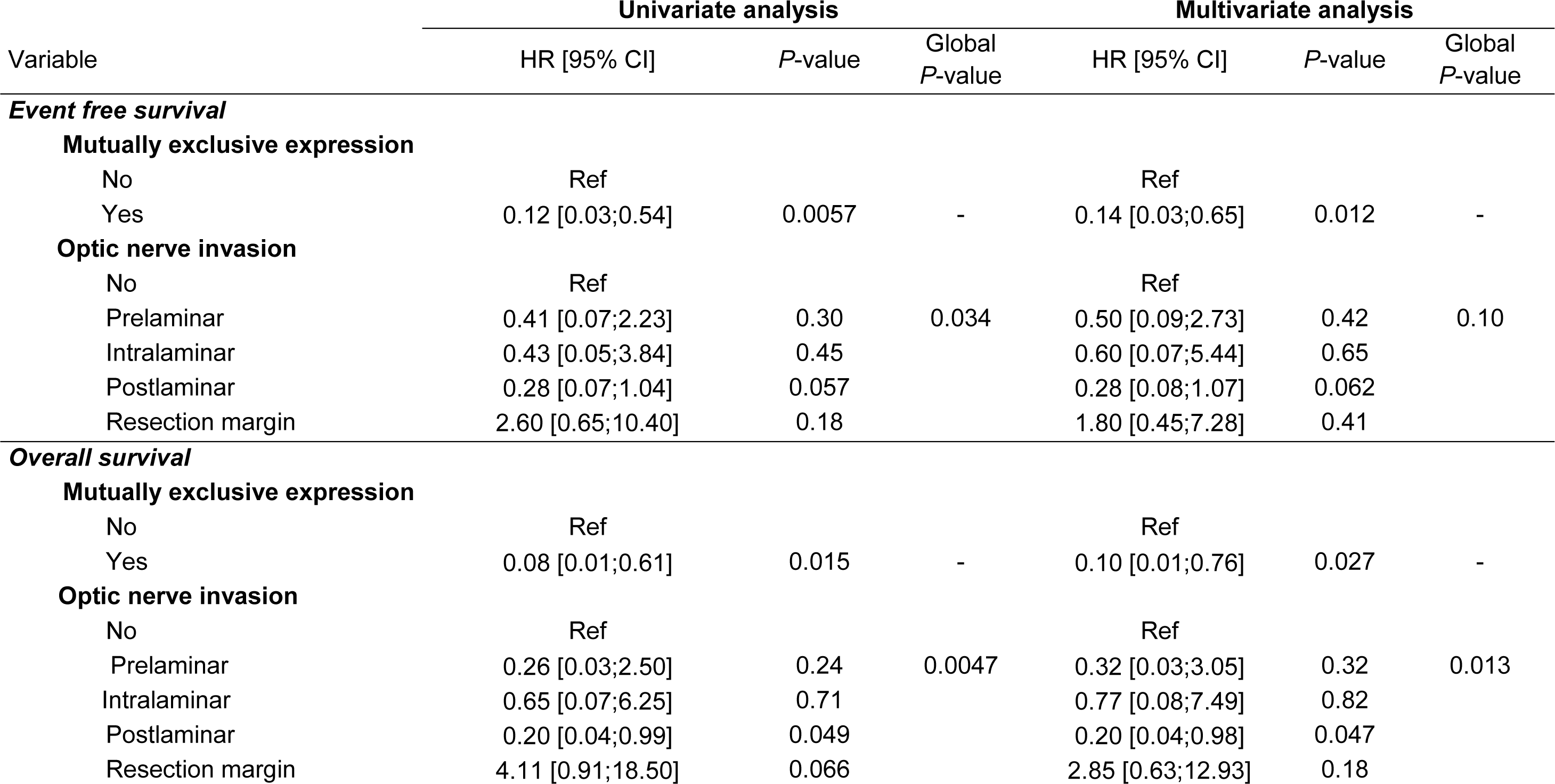
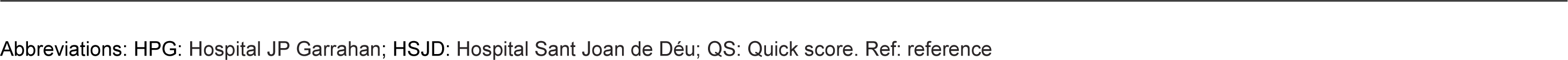
Cox proportional hazards regression models in retinoblastoma patients for association of HS2 tumors with different pattern of expression with probability of event-free survival and overall survival.

| Variable | Univariate analysis |  |  | Multivariate analysis |  |  |
| --- | --- | --- | --- | --- | --- | --- |
|  | HR [95% CI] | P-value | Global P-value | HR [95% CI] | P-value | Global P-value |
| **Event free survival** |  |  |  |  |  |  |
| **Mutually exclusive expression** |  |  |  |  |  |  |
| No | Ref |  |  | Ref |  |  |
| Yes | 0.12 [0.03;0.54] | 0.0057 | - | 0.14 [0.03;0.65] | 0.012 | - |
| **Optic nerve invasion** |  |  |  |  |  |  |
| No | Ref |  |  | Ref |  |  |
| Prelaminar | 0.41 [0.07;2.23] | 0.30 | 0.034 | 0.50 [0.09;2.73] | 0.42 | 0.10 |
| Intralaminar | 0.43 [0.05;3.84] | 0.45 |  | 0.60 [0.07;5.44] | 0.65 |  |
| Postlaminar | 0.28 [0.07;1.04] | 0.057 |  | 0.28 [0.08;1.07] | 0.062 |  |
| Resection margin | 2.60 [0.65;10.40] | 0.18 |  | 1.80 [0.45;7.28] | 0.41 |  |
| **Overall survival** |  |  |  |  |  |  |
| **Mutually exclusive expression** |  |  |  |  |  |  |
| No | Ref |  |  | Ref |  |  |
| Yes | 0.08 [0.01;0.61] | 0.015 | - | 0.10 [0.01;0.76] | 0.027 | - |
| **Optic nerve invasion** |  |  |  |  |  |  |
| No | Ref |  |  | Ref |  |  |
| Prelaminar | 0.26 [0.03;2.50] | 0.24 | 0.0047 | 0.32 [0.03;3.05] | 0.32 | 0.013 |
| Intralaminar | 0.65 [0.07;6.25] | 0.71 |  | 0.77 [0.08;7.49] | 0.82 |  |
| Postlaminar | 0.20 [0.04;0.99] | 0.049 |  | 0.20 [0.04;0.98] | 0.047 |  |
| Resection margin | 4.11 [0.91;18.50] | 0.066 |  | 2.85 [0.63;12.93] | 0.18 |  |

349 Abbreviations: HPG: Hospital JP Garrahan; HSJD: Hospital Sant Joan de Déu; QS: Quick score. Ref: reference

## DISCUSSION

Our results in a large patient cohort, show the feasibility and clinical relevance of an immunohistochemical panel as a surrogate of the recently reported retinoblastoma molecular subtypes. We provide additional characterization of patients with HS2 tumors presenting more commonly with HRPF, older age, higher likelihood of metastatic dissemination at diagnosis and statistically significant higher risk of metastatic relapse and death of disease. Furthermore, we found a higher-risk subgroup among patients with HS2 tumors, identifiable by the presence of a non-mutually exclusive histopathological expression of TFF1 and ARR3. Because immunohistochemistry is available at relatively low cost in many centers treating retinoblastoma worldwide, it would be feasible in clinical scenarios with limited resources for genomic studies.

TFF1 was selected for our study based on its high expression in subtype 2 retinoblastoma in a multi-omic study and its role as a biomarker in retinoblastoma has recently been proposed^9^. Other investigators had previously described a relationship of TFF1 expression with higher TNM clinical stage^21^ and as predictive marker of advanced stage in aqueous humor biopsies^15^.

Many studies have tried to identify biological risk factors in retinoblastoma, but most correlated putative factors with the presence of HRPF and not to actual occurrence of extraocular relapse and/or death^22^. Although nearly all patients who develop an extraocular relapse have HRPF, more than 90% of the patients with HRPF do not relapse and many even without other treatment than enucleation. Therefore, the correlation of any biological feature with the presence of HRPF has a low probability to identify the true high-risk patient who is the one with a high-risk for extraocular relapse despite standard treatment and is therefore at risk of dying.

Our data show a significant difference in EFS and OS between patients with TFF1 positive *versus* negative immunohistochemical staining. This information may be helpful for taking future clinical decisions since the risk of extraocular relapse for patients with HS1 was limited to those declining timely enucleation, regardless of HRPF. All patients with intraocular HS1 retinoblastoma achieved long-term survival, indicating that de-escalation of therapy is likely to be safe in these patients. This is in line with the reported observation that subtype 1 tumors show a stable genome with very few genomic events besides *RB1* alterations^9^. Since subtype 1 is more common in patients with germline *RB1* mutations, the avoidance of adjuvant therapy in these patients is critical to prevent long-term chemo and radiotherapy side effects that including potentially fatal treatment-related secondary malignancies^23^.

All cases presenting with metastatic disease and all those with extraocular relapse (refusal of enucleation excluded) had HS2 tumors, which may be justified by their higher genomic instability and presence of additional chromosomal alterations and mutations. Our data showed a significantly higher expression of TFF1 when intraocular cases with no HRPF were compared to those with HRPF and when those with HRPF with no metastasis are compared to those who developed metastasis. However, it was not possible to find a cut-off value for assigning a very high-risk group based only on TFF1 expression and QS was not associated independently to poor prognosis in multivariate analysis. Notably, within the cohort of patients with HS2 tumors, we show that the non-exclusive expression pattern of ARR3/TFF1 was an additional risk factor in the multivariate analysis.

Based on our data, we would suggest that patients with HS2 and HRPF, especially if a non-mutually exclusive pattern is detected are at higher risk of extraocular relapse and should receive adjuvant treatment. In cases with non-mutually exclusive pattern of ARR3/TFF1, a higher intensity adjuvant therapy may be considered in future studies. Genomic studies to detect additional chromosomal abnormalities such as gains in 1p, 17q and 19q, losses of 11q and mutations in *BCOR* and *MYCN* gains/amplification may provide additional information for further risk stratification. Studies of minimal residual disease or more recently, liquid biopsy may also contribute to describe a very high-risk group warranting intensive treatment^24-27^.

Some limitations to our work need to be acknowledged. This study was done on archival specimens over a long period of time and only limited number of slides were available for some cases. In addition, patients received treatment across successive studies and the efficacy of adjuvant therapy may alter the prognostic significance of TFF1 expression. The additional use of other markers such as EBF3, as reported by Liu et al.^9^, or molecular studies may provide additional information. In our series, germline *RB1* mutations were not studied systematically in our cohort, therefore patients with heritable unilateral retinoblastoma may have been missed if no family history was present. Finally, despite the large number of patients included in the study the number of events was relatively low. Prospective validation of these immunohistochemistry criteria together with other biological parameters in multicentric prospective studies are needed for their use in standard treatment. In cases where adjuvant therapy is debated (for instance isolated choroidal invasion or even retrolaminar optic nerve invasion) it could be discussed to incorporate this subtypes classification as a one more factor in favor of the need or withdrawal of adjuvant treatment.

## CONCLUSIONS

TFF1 expression detected by immunohistochemistry as a surrogate for subtype 2 tumors, especially when it presents with a non-mutually exclusive pattern with ARR3, is a new prognostic factor independently associated to higher risk of extraocular relapse and death in patients with retinoblastoma and HRPF.

## Data Availability

All data produced in the present work are contained in the manuscript

## PRÉCIS

TFF1 expression is a prognostic marker of extraocular relapse and death in patients with high-risk retinoblastoma.

## ACKNOWLEDGMENTS (FINANCIAL SUPPORT)

This research was funded by the Agencia Nacional de Promoción Científica y Tecnológica (Buenos Aires, Argentina) (PIDC 2014-0043); Fondation Nelia et Amadeo Barletta (Nyon, Switzerland); Ligue Nationale Contre le Cancer and Rétinostop (Paris, France); Fundación Leo Messi (Barcelona, Spain); Fund for Ophthalmic Knowledge (New York, United States of America) Instituto Nacional del Cáncer (Buenos Aires, Argentina); Fundación Natali Dafne Flexer de Ayuda al Niño con Cancer (Buenos Aires, Argentina); Fundación Garrahan (Buenos Aires, Argentina) and Instituto Oncológico Henry Moore (Buenos Aires, Argentina).

The sponsors or funding organizations had no role in the design or conduct of this research.

## ABBREVIATIONS/ACRONYMS

ARR3: Arrestin-C
CI: Confidence interval
CNA: Copy number alterations
CNS: Central nervous system
CRX: Cone-rod homeobox
EFS: Event-free survival
HPG: Hospital de Pediatría JP Garrahan
HR: Hazard ratio
HRPF: High-risk pathology factors
HS1: Histological subtype 1
HS2: Histological subtype 2
INCA: Instituto Nacional de Cáncer
IRSS: International retinoblastoma stage system
MSKCC: Memorial Sloan Kettering Cancer Center
OS: Overall survival
QS: Quick score
SJD: Hospital Sant Joan de Déu
TFF1: Trefoil factor 1

## AUTHOR CONTRIBUTIONS

**Conceptualization,** R.A, F.R, P.S, F.L, G.C; **Data curation,** D.G and S.P.J; **Formal analysis:** R.A, D.G and S.P.J; **Funding acquisition**, P.S and G.C; **Investigation**, R.A, G.L, C.R.P, D.O, S.Z, S.C, E.N, M.C.V, G.P.P, C.S, N.G, B.S, M.S, A.M.C; **Methodology**, R.A, D.G, S.P.J; S.C, E.N; **Project administration,** G.C; **Resources,** A.M.C, F.R, D.H.A, P.S, F.L and G.C; **Software**, D.G, S.P.J; **Supervision**, G.C; **Visualization,** R.A, D.G, G.L and S.P.J; **Writing—original draft preparation**, R.A and G.C; **Writing—review and editing**, A.M.C, J.M, M.T.G.D, F.D, F.R, D.H.A, A.S.L, P.S and G.C;

All authors have read and agreed to the published version of the manuscript.

