## Supplemental material for "Immunohistochemical expression of TFF1 is a new prognostic marker in retinoblastoma"

### SUPPLEMENTARY CONTENT

**Figure S1.** Patients classification according to International Retinoblastoma Staging System (IRSS).

**Figure S2.** Clinical features of patients with disseminated retinoblastoma.

**Table S1.** Primary antibodies and the conditions of their use for immunostaining.

**Table S2.** Excel file attached: Samples included in the analyses.

**Table S3.** Clinical characteristics of patients included the in study.

**Table S4.** Histopathological characteristics of patients included in the study.

**Table S5**. Univariate analysis of event-free survival in retinoblastoma patients classified according to the histological subtype.

**Table S6.** Univariate analysis of overall survival in retinoblastoma patients classified according to the histological subtype.

**Table S7**. Univariate analysis of event-free survival in HS2 retinoblastoma patients classified as tumor with mutually or non-mutually exclusive expression pattern.

**Table S8.** Univariate analysis of overall survival in HS2 retinoblastoma patients classified as tumor with mutually or non-mutually exclusive expression pattern.

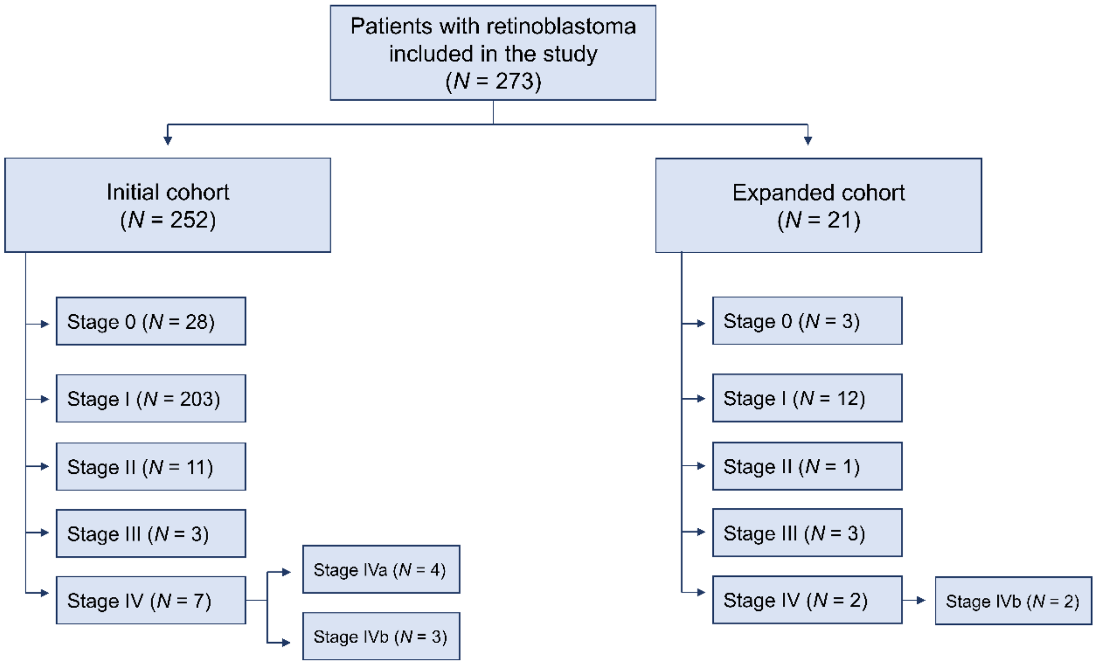


**Figure S1. Patients classification according to International Retinoblastoma Staging System (IRSS).** Flow chart and IRSS classification of patients included in the initial and extended cohort.

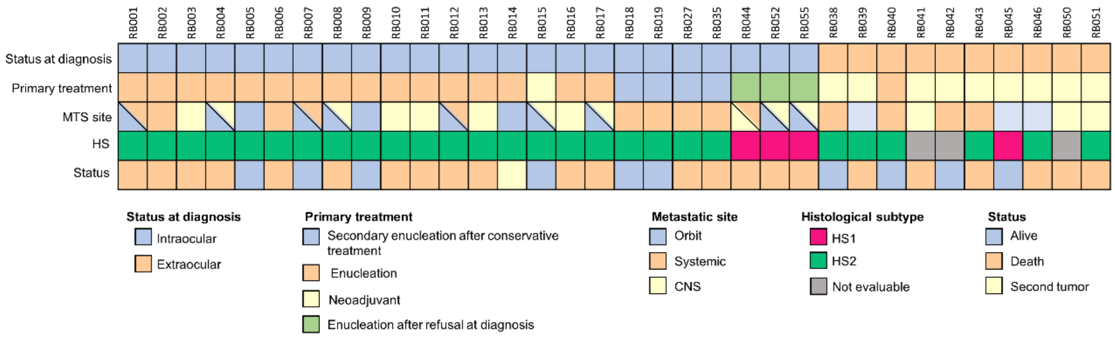


**Figure S2. Clinical features of patients with disseminated retinoblastoma.** Each column represents an individual sample, and the boxes correspond to the status of each of the main characteristics depicted. Boxes are partiotioned if more than one relevant feature coexists.

**Table S1. Primary antibodies and the conditions of their use for immunostaining.**

| Antibody | Antigen retrieval | Working dilution | Exposition (Time - temperature) | Positive control | Staining pattern |
|---|---|---|---|---|---|
| CRX | pH 6 | 1/500 | 60 minutes - RT | Retina | Nuclear |
| ARR3 | pH 6 | 1/300 | 60 minutes - RT | Retina | Cytoplasmic |
| TFF1 | pH 6 | 1/3000 | 60 minutes - RT | Stomach | Cytoplasmic |
| KI67 | pH 6 | 1/100 | 60 minutes - RT | Medulloblastoma tumor | Nuclear |

Abbreviations: RT: Room temperature.

**Table S3. Clinical characteristics of patients included the in study.**

| | Total (*N* = 273) | Stage 0-II (*N* = 258) | Stage III-IV (*N* = 15) |
|---|---|---|---|
| **Center** | | | |
| HPG | 210 (76.9%) | 200 (77.5%) | 10 (66.7%) |
| HSJD | 42 (15.4%) | 42 (16.3%) | 0 (0.0%) |
| INCA | 11 (4.0%) | 8 (3.1%) | 3 (20.0%) |
| MSKCC | 7 (2.6%) | 7 (2.7%) | 0 (0.0%) |
| Hosp. de Niños M. A. Villarroel | 2 (0.7%) | 0 (0.0%) | 2 (13.3%) |
| Hospital Pereira Rossell | 1 (0.4%) | 1 (0.4%) | 0 (0.0%) |
| **Gender** | | | |
| Male | 147 (53.8%) | 139 (53.9%) | 8 (53.3%) |
| Female | 126 (46.2%) | 119 (46.1%) | 7 (46.7%) |
| **Age at diagnosis (months)** | | | |
| Mean (SD) | 29.4 (22.7) | 29.1 (23.0) | 34.7 (16.2) |
| Median [Min, Max] | 27.0 [0.0, 166.0] | 27.0 [0.0, 166.0] | 34.0 [15.0, 76.0] |
| **Laterality** | | | |
| Unilateral | 205 (75.1%) | 193 (74.8%) | 12 (80.0%) |
| Bilateral | 68 (24.9%) | 65 (25.2%) | 3 (20.0%) |
| **Primary treatment** | | | |
| Enucleation | 221 (81.0%) | 217 (84.1%) | 4 (26.7%) |
| Conservative | 31 (11.4%) | 31 (12.0%) | 0 (0.0%) |
| Neoadjuvant | 16 (5.9%) | 5 (1.9%) | 11 (73.3%) |
| Enucleation refusal | 5 (1.8%) | 5 (1.9%) | 0 (0.0%) |
| **Adjuvant therapy** | | | |
| Yes | 176 (64.5%) | 162 (62.8%) | 14 (93.3%) |
| No | 97 (35.5%) | 96 (37.2%) | 1 (6.7%) |
| **Relapse** | | | |
| Yes | 48 (17.6%) | 40 (15.5%) | 8 (53.3%) |
| No | 225 (82.4%) | 218 (84.5%) | 7 (46.7%) |
| **Outcome** | | | |
| Alive disease free | 236 (86.4%) | 231 (89.5%) | 5 (33.3%) |
| Death | 35 (12.8%) | 26 (10.1%) | 9 (60.0%) |
| Second tumor development | 1 (0.4%) | 0 (0.0%) | 1 (6.7%) |
| Lost to follow up | 1 (0.4%) | 1 (0.4%) | 0 (0.0%) |

Abbreviations: HPG: Hospital de Pediatría Garrahan, HSJD: Hospital Sant Joan de Déu, INCA: Instituto Nacional de Câncer, MSKCC: Memorial Sloan Kettering Cancer Center.

**Table S4.** Histopathological characteristics of patients included in the study.

| | **Total (*N* = 273)** | **Stage 0-II (*N* = 258)** | **Stage III-IV (*N* = 15)** |
|---|---|---|---|
| **HRPF** | | | |
| Yes | 181 (66.3%) | 173 (67.1%) | 8 (53.3%) |
| No | 84 (30.8%) | 84 (32.6%) | 0 (0.0%) |
| Not evaluable | 6 (2.2%) | 1 (0.4%) | 5 (33.3%) |
| Not enucleated | 2 (0.7%) | 0 (0.0%) | 2 (13.3%) |
| **pTNM** | | | |
| pT1 | 28 (10.3%) | 28 (10.9%) | 0 (0.0%) |
| pT2 | 56 (20.5%) | 56 (21.7%) | 0 (0.0%) |
| pT3a | 33 (12.1%) | 33 (12.8%) | 0 (0.0%) |
| pT3b | 78 (28.6%) | 76 (29.5%) | 2 (13.3%) |
| pT3c | 46 (16.8%) | 46 (17.8%) | 0 (0.0%) |
| pT3d | 7 (2.6%) | 5 (1.9%) | 2 (13.3%) |
| pT4 | 17 (6.2%) | 13 (5.0%) | 4 (26.7%) |
| pTX* | 6 (2.2%) | 1 (0.4%) | 5 (33.3%) |
| Not enucleated | 2 (0.7%) | 0 (0.0%) | 2 (13.3%) |

* Not evaluable due to necrosis higher than 90% of tumor.
Abbreviations: pT: pathological primary tumor. HRPF: high-risk pathology factors.

**Table S5**. Univariate analysis of patients with intraocular retinoblastoma included in the initial cohort for association of clinical and histopathological features and histological subtypes classification with event-free survival.

| Variable | HR [95% CI] | *P*-value | Global *P*-value |
|---|---|---|---|
| **Centre** | | | |
| HPG | Ref | | |
| HSJD | 0.52 [0.12;2.24] | 0.38 | - |
| **Gender** | | | |
| Female | Ref | | |
| Male | 1.06 [0.43;2.61] | 0.90 | - |
| **Initially enucleated** | | | |
| No | | | |
| Yes | 0.59 [0.20;1.79] | 0.35 | - |
| **Laterality** | | | |
| Bilateral | | | |
| Unilateral | 0.78 [0.30;2.04] | 0.61 | - |
| **Age at diagnosis** | | | |
| < 9 months | Ref | | |
| 9 - 18 months | 0.73 [0.07;8.01] | 0.79 | 0.14 |
| > 18 months | 2.68 [0.62;11.70] | 0.19 | |
| **Adjuvant therapy** | | | |
| No | Ref | | |
| Yes | 1.55 [0.56;4.31] | 0.40 | - |
| **Choroid invasion** | | | |
| No | Ref | | |
| Focal | 0.93 [0.21;4.17] | 0.93 | 0.59 |
| Massive | 1.55 [0.44;5.48] | 0.50 | |
| **Optic nerve invasion** | | | |
| No | Ref | | |
| Prelaminar | 0.39 [0.08;1.93] | 0.25 | 0.0036 |
| Intralaminar | 0.52 [0.06;4.29] | 0.54 | |
| Postlaminar | 0.43 [0.13;1.42] | 0.17 | |
| Resection margin | 6.07 [1.84;20.0] | 0.0031 | |
| **Sclera invasion** | | | |
| No | Ref | | |
| Intrascleral | 1.25 [0.40;3.89] | 0.69 | 0.083 |
| Transcleral | 5.63 [1.59;20.00] | 0.0075 | |
| **Histological subtype** | | | |
| HS1 | Ref | | |
| HS2 | 28.50 [3.92;3626.36] | < 0.0001 | - |

Patient who developed secondary neoplasm was excluded in the analysis. Abbreviations: HPG: Hospital JP Garrahan; HSJD: Hospital Sant Joan de Déu; HS1: Histological subtype 1; HS2: Histological subtype 2.

**Table S6**. Univariate analysis in patients with intraocular retinoblastoma included in the initial cohort for association of clinical and histopathological features and histological subtypes classification with overall survival.

| Variable | HR [95% CI] | *P*-value | Global *P*-value |
|---|---|---|---|
| **Centre** | | | |
| HPG | Ref | | |
| HSJD | 0.33 [0.04;2.55] | 0.29 | - |
| **Gender** | | | |
| Female | Ref | | |
| Male | 0.95 [0.33;2.72] | 0.93 | - |
| **Initially enucleated** | | | |
| No | Ref | | |
| Yes | 0.92 [0.21;4.12] | 0.91 | - |
| **Laterality** | | | |
| Bilateral | Ref | | |
| Unilateral | 0.91 [0.29;2.91] | 0.88 | - |
| **Age at diagnosis** | | | |
| < 9 months | Ref | | |
| 9 - 18 months | 1.50 [0.09;24.00] | 0.74 | 0.18 |
| > 18 months | 4.02 [0.52;31.00] | 0.18 | |
| **Adjuvant therapy** | | | |
| No | Ref | | |
| Yes | 1.30 [0.41;4.17] | 0.66 | - |
| **Choroid invasion** | | | |
| No | Ref | | |
| Focal | 0.21 [0.02;2.07] | 0.18 | 0.12 |
| Massive | 1.20 [0.33;4.38] | 0.78 | |
| **Optic nerve invasion** | | | |
| No | Ref | | |
| Prelaminar | 0.29 [0.03;2.59] | 0.27 | 0.00041 |
| Intralaminar | 0.82 [0.09;7.32] | 0.86 | |
| Postlaminar | 0.36 [0.08;1.63] | 0.19 | |
| Resection margin | 10.4 [2.76;39.00] | 0.00054 | |
| **Sclera invasion** | | | |
| No | Ref | | |
| Intrascleral | 1.41 [0.37;5.30] | 0.61 | 0.026 |
| Transcleral | 9.44 [2.50;35.70] | 0.00093 | |
| **Histological subtype** | | | |
| HS1 | Ref | | |
| HS2 | 20.45 [2.74;2613.57] | 0.00054 | - |

Patient who developed secondary neoplasm was excluded from the analysis. Abbreviations: HPG: Hospital JP Garrahan; HSJD: Hospital Sant Joan de Déu; HS1: Histological subtype 1; HS2: Histological subtype 2.

**Table S7**. Univariate analysis in patients with intraocular retinoblastoma included in the initial cohort for association of presence or absence of mutually exclusive expression pattern and clinical and histopathological characteristics with event-free survival.

| Variable | HR [95% CI] | *P*-value | Global *P*-value |
|---|---|---|---|
| **Centre** | | | |
| HPG | Ref | | |
| HSJD | 0.58 [0.00;4.32] | 0.68 | - |
| **Gender** | | | |
| Female | Ref | | |
| Male | 1.19 [0.45;3.17] | 0.73 | - |
| **Initially enucleated** | | | |
| No | Ref | | |
| Yes | 0.47 [0.11;2.07] | 0.32 | - |
| **Laterality** | | | |
| Bilateral | Ref | | |
| Unilateral | 0.59 [0.20;1.69] | 0.33 | - |
| **Age at diagnosis** | | | |
| < 9 months | Ref | | |
| 9 - 18 months | 0.59 [0.04;9.45] | 0.71 | 0.71 |
| > 18 months | 1.25 [0.16;9.51] | 0.83 | |
| **Adjuvant therapy** | | | |
| No | Ref | | |
| Yes | 1.26 [0.41;3.91] | 0.69 | - |
| **Choroid invasion** | | | |
| No | Ref | | |
| Focal | 0.76 [0.14;4.16] | 0.75 | 0.92 |
| Massive | 0.95 [0.21;4.35] | 0.95 | |
| **Optic nerve invasion** | | | |
| No | Ref | | |
| Prelaminar | 0.41 [0.07;2.23] | 0.30 | 0.034 |
| Intralaminar | 0.43 [0.05;3.84] | 0.45 | |
| Postlaminar | 0.28 [0.07;1.04] | 0.057 | |
| Resection margin | 2.60 [0.65;10.40] | 0.18 | |
| **Sclera invasion** | | | |
| No | Ref | | |
| Intrascleral | 0.60 [0.13;2.73] | 0.51 | 0.14 |
| Transcleral | 3.63 [1.01;13.00] | 0.048 | |
| **TFF1 QS** | | | |
| Low (50 -10) | Ref | | |
| Moderate (101 - 200) | 3.14 [0.38;26.10] | 0.29 | 0.45 |
| High (201-300) | 2.90 [0.37;22.90] | 0.31 | |
| **Mutually exclusive expression** | | | |
| No | Ref | | |
| Yes | 0.12 [0.03;0.54] | 0.0057 | - |

Patient who developed secondary neoplasm was excluded in the analysis. Abbreviations: HPG: Hospital JP Garrahan; HSJD: Hospital Sant Joan de Déu.

**Table S8**. Univariate analysis in patients with intraocular retinoblastoma included in the initial cohort for association of presence or absence of mutually exclusive expression pattern and clinical and histopathological characteristics with overall survival.

| Variable | HR [95% CI] | *P*-value | Global *P*-value |
|---|---|---|---|
| **Centre** | | | |
| HPG | Ref | | |
| HSJD | 0.74 [0.00;5.65] | 0.83 | - |
| **Gender** | | | |
| Female | Ref | | |
| Male | 0.85 [0.27;2.68] | 0.78 | - |
| **Initially enucleated** | | | |
| No | Ref | | |
| Yes | 0.73 [0.009;5.68] | 0.76 | - |
| **Laterality** | | | |
| Bilateral | Ref | | |
| Unilateral | 0.53 [0.16;1.76] | 0.30 | - |
| **Age at diagnosis** | | | |
| < 9 months | Ref | | |
| 9 - 18 months | 0.57 [0.04;9.20] | 0.70 | 0.91 |
| > 18 months | 0.85 [0.11;6.64] | 0.88 | |
| **Adjuvant therapy** | | | |
| No | Ref | | |
| Yes | 1.19 [0.32;4.40] | 0.80 | - |
| **Choroid invasion** | | | |
| No | Ref | | |
| Focal | 0.16 [0.01;1.82] | 0.14 | 0.14 |
| Massive | 0.82 [0.18;3.87] | 0.80 | |
| **Optic nerve invasion** | | | |
| No | Ref | | |
| Prelaminar | 0.26 [0.03;2.50] | 0.24 | 0.0047 |
| Intralaminar | 0.65 [0.07;6.25] | 0.71 | |
| Postlaminar | 0.20 [0.04;0.99] | 0.049 | |
| Resection margin | 4.11 [0.91;18.50] | 0.066 | |
| **Sclera invasion** | | | |
| No | Ref | | |
| Intrascleral | 1.00 [0.21;4.83] | 1.00 | 0.056 |
| Transcleral | 6.84 [1.77;26.50] | 0.0054 | |
| **TFF1 QS** | | | |
| Low (50 -10) | Ref | | |
| Moderate (101 - 200) | 2.78 [0.33;23.80] | 0.35 | 0.58 |
| High (201-300) | 1.96 [0.24;16.30] | 0.53 | |
| **Mutually exclusive expression** | | | |
| No | Ref | | |
| Yes | 0.08 [0.01;0.61] | 0.015 | - |

Patient who developed secondary neoplasm was excluded from the analysis. Abbreviations: HPG: Hospital JP Garrahan; HSJD: Hospital Sant Joan de Déu.
